# Seasonal variation in blood pressure recorded in routine primary care

**DOI:** 10.1101/2021.11.15.21266359

**Authors:** Armindokht Shahsanai, Sumeet Kalia, Babak Aliarzadeh, Rahim Moineddin, Aashka Bhatt, Michelle Greiver

## Abstract

**Objective:** Seasonal variations in blood pressure (BP) exist. There is limited information about important clinical factors associated with increased BP and the strength and amplitude of seasonal variation in primary care.

**Methods:** This was a repeated cross-sectional observational study of routinely measured BPs in primary care using data from electronic medical records in the greater Toronto region, from January 2009 to June 2019. We used time-series models and mean monthly systolic BPs (SBPs) and diastolic BPs (DBPs) to estimate the strength and amplitude of seasonal oscillations, as well as their associations with patient characteristics.

**Results:** 314,518 patients were included. Mean SBPs and DBPs were higher in winter than summer. There was strong or perfect seasonality for all characteristics studied, except for BMI less than 18.5 (underweight). Overall, the mean maximal amplitude of the oscillation was 1.51mmHg for SPB (95% CI 1.30mmHg to 1.72mmHg) and 0.59mmHg for DBP (95% CI 0.44mmHg to 0.74mmHg). Patients aged 81 years or older had larger SBP oscillations than younger patients aged 18 to 30 years; the difference was 1.20mmHg (95% CI 1.15mmHg to 1.66mmHg). Hypertension was also associated with greater oscillations, difference 0.53mmHg (95% CI 0.18mmHg to 0.88mmHg). There were no significant differences in SBP oscillations by other patient characteristics, and none for DBP.

**Conclusion:** Strong seasonality was detected for almost all patient subgroups studied and was greatest for older patients and for those with hypertension. The variation in BP between summer and winter should be considered by clinicians when making BP treatment decisions.

## Introduction

Managing blood pressure (BP) is one of the most important strategies to prevent future onset of cardiovascular disease (1-3). Hypertension is a known precursor for cardiovascular events such as myocardial infraction, stroke or premature death (4).

Although clinical guidelines are designed to emphasize the importance of lowering BP to reduce the future onset of cardiovascular-related adverse events(5), it remains unclear as to how these clinical guidelines account for known seasonal variations of systolic and diastolic blood pressures (SBP; DBP).

Many factors, including physiological (6) and environmental (7) are known to influence the seasonal variation of BP. Previous studies have documented seasonal BP fluctuations, with peaks recorded during winter months and troughs recorded during the summer months (7-16). These seasonal fluctuations are observed with different modes of recording BP including clinical office-based measurements, self-recorded measurements and 24-hour ambulatory BP monitoring (17). Ambient temperature is one of the main environmental factor associated with seasonal variations in BP. (17-24).Other environmental factors such as altitude, latitude, air pollutants (7) and daylight hours (16) are also known to influence BP regulation.

Apart from environmental factors, the amount of fluctuation in seasonal BP may also vary with respect to patient characteristics. For example, Lewigton et al. reported increased variation in BP among elderly patients. (19) While other studies demonstrated increased variation in seasonal BP among hypertensive patients (25-27). Cois and Ehrlich showed that seasonal variations of BP were higher among subjects in the lowest socioeconomic classes than in the highest (28). Kristal-Boneh et al. noted that BP variations from summer to winter are inversely associated with body mass index and weight (29). The magnitude of these oscillations in BP with respect to important patient characteristics may further have clinical implications as they can affect treatment decisions. For instance, Sakamoto et al indicated achievement rates for guideline targets for blood pressure were greater in the summer than in the winter for people living with diabetes (30).

Hypertension is most often diagnosed and managed in primary care, making this an ideal setting to study real life seasonal variations and associations with patient characteristics. The objectives of this study are: 1) to quantify the strength of BP seasonality with respect to demographic, phenotypic and socioeconomic characteristics; 2) to quantify the amplitude of seasonal fluctuation in monthly BP among adult patients in primary care practices.

## Methods

This is a repeated cross-sectional observational study design using BP measured in routine visits to primary care practices between January 1, 2009 and June 30, 2019. We applied the STrengthening the Reporting of OBservational studies in Epidemiology checklist for this observational study (31).

### Settings and data sources

The Canadian Primary Care Sentinel Surveillance Network (CPCSSN) is Canada’s largest electronic medical record (EMR)-based chronic disease surveillance system (32). De-identified patient information is collected from primary care practice-based research networks across Canada; the data are aggregated in a single, secure central database housed in Kingston, Ontario (33). In this study, we used data from the University of Toronto Practice-Based Research Network (UTOPIAN). UTOPIAN is the largest network in CPCSSN, with one-third of data in the national database; it includes providers and patients from Toronto and surrounding areas in southern Ontario, Canada.(34) We used data extracted on or after June 30^th^ 2019.

### Study population and variables

We used an open cohort to enroll patients from Jan 2009 to Jun 2019 using the following inclusion criteria: (1) patient registered to participating practices; (ii) patient is at least 18 years of age at the time of blood pressure measurement. The data elements in the final cohort included age (as of the date of each BP measurement), sex, income quintiles, region (urban/rural), SBP, DBP, measurement date, body mass index (BMI), presence of hypertension and presence of diabetes. The urban/rural classification was computed using the second digit of Canada’s residential postal codes (which contains six elements) where 0=rural; non-zero=urban (35). The definition for hypertension or diabetes was derived from the validated case definition for these conditions in CPCSSN.(36) BMI, hypertension and diabetes were defined as time-dependent covariates over the course of study follow-up (Jan 01, 2009 to Jun 30, 2019). Since the information pertaining to BMI and diagnosis of hypertension and diabetes can be recorded at multiple time points during the study follow-up from January 2009 to Jun 2019, time-dependent covariates were used based on the most recent observation available at the time when the BP was measured. In particular, the earliest diagnosis of a chronic conditions was carried forward for the remainder of the study follow-up for a given patient; in other words, once a patient was diagnosed as having diabetes, they are labeled as having this condition for all observations going forward.

The study was reviewed and approved by the University of Toronto’s Research Ethics Board. The dataset was curated using SQL and statistical analysis were performed using SAS v.9.4.

### Statistical Analysis

We calculated mean SBP and DBP for each month from January 2009 to June 2019 and used this to calculate the strength and amplitude of seasonal variations. The amplitude was measured by the difference between the month with highest mean BP and the month with lowest mean BP.

The coefficient of determination of autoregressive model (R^2^_Autoreg_)was used to quantify the strength of seasonality. The magnitude of R^2^_Autoreg_ lies between 0 and 1 where zero indicates no seasonality, and one indicates perfect seasonality. The strength of seasonality is based on values of R^2^_Autoreg_, where 0 to less than 0.1 represents non-existent seasonality; 0.1 to 0.4 represents weak seasonality; 0.4 to less than 0.6 represents moderate seasonality; 0.6 to less than 0.9 represents strong seasonality and 0.9 to 1 represent to perfect seasonality(37). We also applied an alternative test for seasonality using Bartlett’s Kolmogorov-Smirmov (BKS) test. The BKS test is based on the comparison of the normalized cumulative periodogram with the cumulative distribution function of a uniform zero and one random variable (37).

We modeled the mean SBP and DBP using spectral analysis. The sinusoidal components were used to estimate the oscillation in mean SBP and DBP in a smooth periodic fashion where the trend was taken into account as the duration of study follow-up using each calendar year. The following time-series models were used for mean SBP and DBP with respect to each sub-group defined based on patient characteristics:

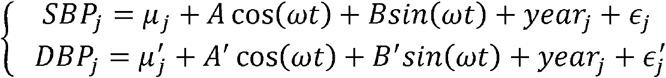

Where *μ*_*j*_ represents the baseline mean for characteristic *j, A* = *α* sin(*δ*); *B* = *α* cos(*δ*) and 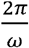 correspond to the complete period of sinusoid function(38). The parameter represents the amplitude of sinusoid function; this can be recovered from the estimated values of regression coefficients as 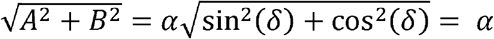. The amplitude is defined as the difference between the cyclic peaks and cyclic troughs evident in the sinusoid curves. The phase shift and *δ* the frequency of periodicity *ω* are both measured in radians and *ε*_*j*_ is the random error component assumed to be distributed as *N*(0, σ^2^).

## Results

The open cohort included 314,518 patients by the end of study follow-up (June 30, 2019). Patient characteristics are shown in Table 1. The mean SBP was higher in older patients, in males, in those with higher BMIs and in those with diabetes or hypertension.

**Table 1:** Mean systolic and diastolic blood pressure with respect to patient characteristics.

|  | Total number of patients* |  | Systolic blood pressure |  | Diastolic blood pressure |  |
| --- | --- | --- | --- | --- | --- | --- |
|  | N* | Column % | Mean | std dev | Mean | std dev |
| **Age group** |  |  |  |  |  |  |
| 18-30 years | 45101 | 14.34% | 114.47 | 14.24 | 72.49 | 10.25 |
| 31-40 years | 50887 | 16.18% | 117.55 | 16.13 | 75.59 | 11.46 |
| 41-50 years | 53451 | 16.99% | 123.81 | 17.87 | 79.48 | 11.50 |
| 51-60 years | 60204 | 19.14% | 128.10 | 18.35 | 79.76 | 10.80 |
| 61-70 years | 49465 | 15.73% | 130.53 | 18.88 | 77.26 | 10.48 |
| 71-80 years | 33161 | 10.54% | 131.77 | 19.43 | 74.00 | 10.50 |
| 81+ years | 22249 | 7.07% | 132.81 | 21.12 | 71.41 | 10.93 |
| **Sex** |  |  |  |  |  |  |
| Female | 182236 | 57.94% | 125.30 | 19.68 | 75.48 | 11.03 |
| Male | 132282 | 42.06% | 128.50 | 18.26 | 77.91 | 11.34 |
| **Region** |  |  |  |  |  |  |
| Missing | 4257 | 1.35% | 127.80 | 20.34 | 75.74 | 11.28 |
| Rural | 26507 | 8.43% | 129.40 | 18.22 | 77.09 | 11.06 |
| Urban | 283754 | 90.22% | 126.26 | 19.25 | 76.38 | 11.23 |
| **Income Quintiles** |  |  |  |  |  |  |
| Missing | 37882 | 12.04% | 125.02 | 17.75 | 76.32 | 10.72 |
| 1(=lowest) | 49853 | 15.85% | 127.45 | 19.33 | 76.12 | 11.19 |
| 2 | 46733 | 14.86% | 127.33 | 19.43 | 76.25 | 11.20 |
| 3 | 47038 | 14.96% | 127.06 | 19.23 | 76.49 | 11.18 |
| 4 | 54577 | 17.35% | 126.25 | 19.34 | 76.88 | 11.46 |
| 5(=highest) | 78435 | 24.94% | 126.02 | 19.49 | 76.50 | 11.34 |
| **Body Mass index (Kg/m<sup>2</sup>)** |  |  |  |  |  |  |
| Missing | 73217 | 23.28% | 125.65 | 20.01 | 77.06 | 11.78 |
| 18.4 or less (Underweight) | 4763 | 1.51% | 115.32 | 19.58 | 70.43 | 10.50 |
| 18.5 - 24.9 (Normal) | 77870 | 24.76% | 121.42 | 19.02 | 73.17 | 10.47 |
| 25 - 29.9 (overweight) | 84720 | 26.94% | 127.68 | 18.30 | 76.20 | 10.64 |
| >=30 (Obese) | 73948 | 23.51% | 130.75 | 17.69 | 78.12 | 10.82 |
| **Hypertension** |  |  |  |  |  |  |
| No | 235729 | 74.95% | 121.54 | 17.37 | 75.27 | 10.67 |
| Yes | 78789 | 25.05% | 133.45 | 19.45 | 78.03 | 11.74 |
| **Type II Diabetes** |  |  |  |  |  |  |
| No | 275804 | 87.69% | 125.56 | 19.22 | 76.71 | 11.25 |
| Yes | 38714 | 12.31% | 130.13 | 18.70 | 75.41 | 11.04 |
| **Total** | 314518 | 100% | 126.55 | 19.20 | 76.43 | 11.22 |

\*patient count (N) is based on total number of patients at the end of the study follow-up (Jun 2019); we used the most recent information available for body mass index, hypertension and diabetes.

The mean SBP was 124.59mmHg (95% CI 124.51mmHg to 124.67mmHg) in the peak summer month (July) and 127.74mmHg (95% CI 127.66mmHg to 127.84mmHg) in the through winter month (January); mean DBP was 76.93mmHg (95% CI 76.89mmHg to 76.98mmHg) and 75.78mmHg (95% CI 75.73mmHg to 75.82mmHg).

Table 2 provides the strength of seasonality by patient characteristics. There was strong or perfect seasonality for all characteristics studied, except for BMI less than 18.5 (underweight). Figure 1 shows the amplitude of seasonal BP fluctuation. Overall, the mean amplitude of the oscillation was 1.51mmHg for SBP (95% CI 1.30mmHg to 1.72mmHg) and 0.59mmHg for DBP (95% CI 0.44mmHg to 0.74mmHg) in the entire study population. There were significant differences in the amplitude of oscillations for SBP between younger and older patients (0.92mmHg in 18-29 year vs 2.12mmHg in 81+ years), difference 1.20mmHg (95% CI 1.15mmHg to 1.66mmHg) and for those with and without hypertension (1.34mmHg without and 1.87mmHg with, difference 0.53mmHg (95% CI 0.18mmHg to 0.88mmHg). There were no significant differences by patient characteristics for DBP, or for characteristics other than age and hypertension for SBP. Supplementary figures (S1-S7) in the appendix section show the oscillations of both systolic and diastolic blood pressures with respect to age group, sex, BMI, hypertension, diabetes, rurality and income quintiles.

**Table 2:** Strength of seasonality by patient characteristics.

| Variable | Characteristics | Systolic blood pressure |  |  | Diastolic blood pressure |  |  |
| --- | --- | --- | --- | --- | --- | --- | --- |
| | | BKS statistic* | $R^2_{\text{autoreg}}$ | Strength | BKS statistic* | $R^2_{\text{autoreg}}$ | Strength |
| Age group | 18-30 years | 0.747 | 0.918 | Perfect | 0.758 | 0.805 | Strong |
|  | 31-40 years | 0.720 | 0.843 | Strong | 0.744 | 0.826 | Strong |
|  | 41-50 years | 0.752 | 0.879 | Strong | 0.680 | 0.768 | Strong |
|  | 51-60 years | 0.767 | 0.883 | Strong | 0.751 | 0.893 | Strong |
|  | 61-70 years | 0.761 | 0.890 | Strong | 0.717 | 0.899 | Strong |
|  | 71-80 years | 0.723 | 0.820 | Strong | 0.713 | 0.878 | Strong |
|  | 81+ years | 0.700 | 0.778 | Strong | 0.668 | 0.819 | Strong |
| Sex | Female | 0.758 | 0.929 | Perfect | 0.752 | 0.955 | Perfect |
|  | Male | 0.775 | 0.920 | Perfect | 0.731 | 0.933 | Perfect |
| Region | Rural | 0.687 | 0.845 | Strong | 0.646 | 0.758 | Strong |
|  | Urban | 0.774 | 0.936 | Perfect | 0.750 | 0.956 | Perfect |
| Income quintiles | 1 | 0.750 | 0.874 | Strong | 0.647 | 0.777 | Strong |
|  | 2 | 0.743 | 0.863 | Strong | 0.707 | 0.872 | Strong |
|  | 3 | 0.748 | 0.891 | Strong | 0.687 | 0.897 | Strong |
|  | 4 | 0.767 | 0.921 | Perfect | 0.745 | 0.929 | Perfect |
|  | 5 | 0.757 | 0.923 | Perfect | 0.754 | 0.923 | Perfect |
| BMI | 18.4 or less (Underweight) | 0.485 | 0.475 | Moderate | 0.225 | 0.205 | Weak |
|  | 18.5 – 24.9 (Normal) | 0.734 | 0.883 | Strong | 0.635 | 0.757 | Strong |
|  | 25 – 29.9 (overweight) | 0.740 | 0.846 | Strong | 0.666 | 0.808 | Strong |
| | $\geq 30$ (Obese) | 0.724 | 0.799 | Strong | 0.631 | 0.675 | Strong |
| Hypertension | No | 0.755 | 0.921 | Perfect | 0.768 | 0.954 | Perfect |
|  | Yes | 0.755 | 0.880 | Strong | 0.716 | 0.953 | Perfect |
| Type II diabetes | No | 0.762 | 0.930 | Perfect | 0.753 | 0.956 | Perfect |
|  | Yes | 0.762 | 0.882 | Strong | 0.715 | 0.885 | Strong |

\* Bartlett's Kolmogorov-Smirnov (BKS) test statistic.

**Figure 1:**
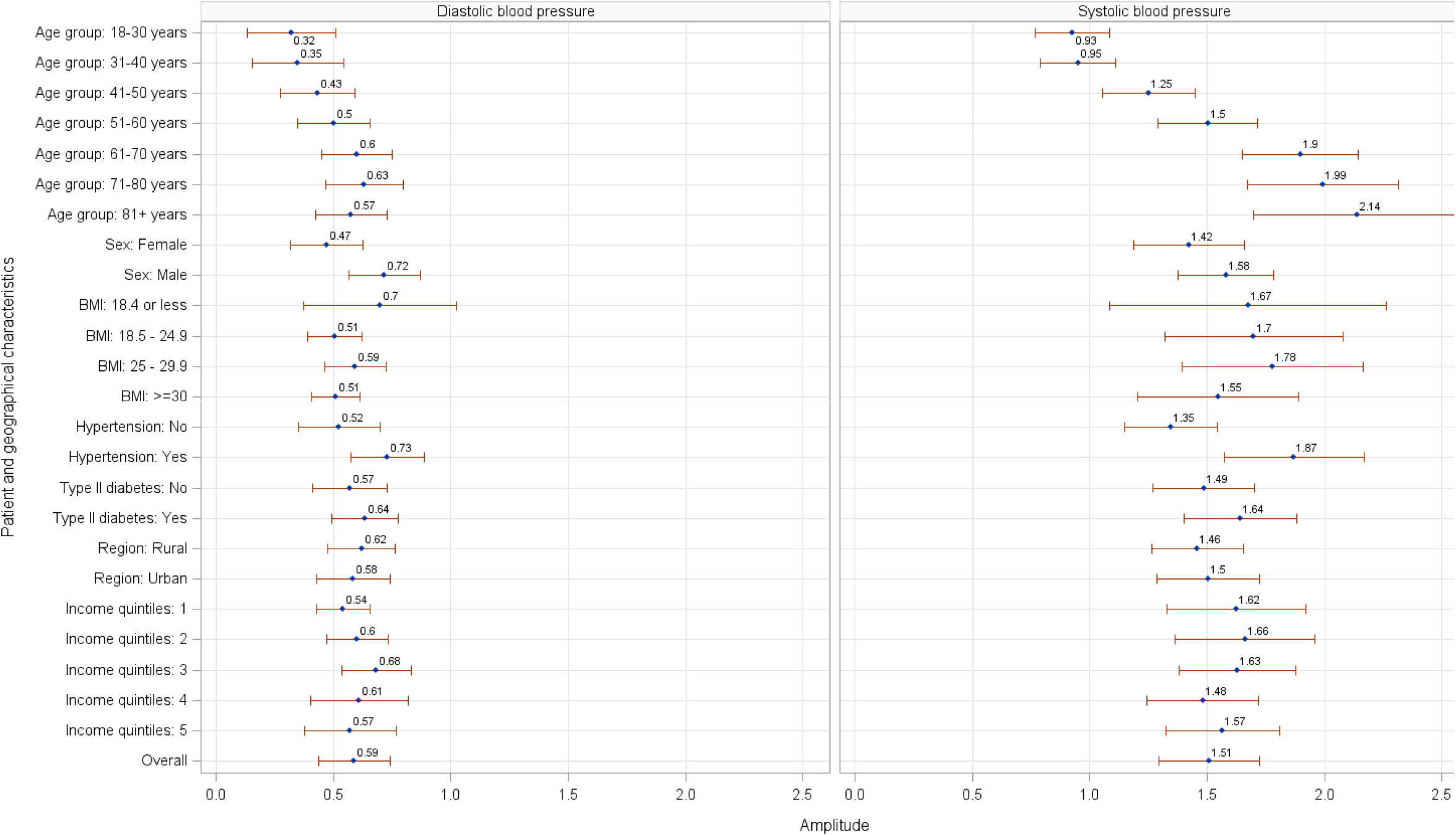
Amplitude of blood pressure variation by patient characteristics.

## Discussion

We found strong seasonal oscillations in BP in this real-world primary care data repository. The difference in amplitude was greatest for the oldest, when compared with the youngest and for those with hypertension, compared to those without.

Several studies have noted seasonal variation in BP (20, 39) including a large study in China(19) and in the Medical Research Council’s treatment trial over 17 000 men and women throughout England, Scotland, and Wales.(13). In agreement with previous studies, the seasonal effect was also noticeable for SBP with respect to different patient characteristics (28, 40).

In this study, we filled an important gap in the literature by quantifying the magnitude of seasonal variation in both SBP and DBP with respect to important patient characteristics. In general, we found a strong seasonal pattern in blood pressure in this primary care adult population, with higher mean BPs in the winter. The pattern was persistent over the decade studied. Strong seasonal variations occurred in almost all patient subgroups (except for underweight BMI group). There were significant differences in the amplitude of seasonal oscillations for SBP between younger and older patients. This finding is in agreement with the results of an earlier study which showed the BP oscillations were greater among elderly participants than in younger participants.(11) Although our study did not find differences in seasonal variation by socioeconomic status, Cois et al showed seasonal effects were lower among subjects in the highest socioeconomic classes than those in the lowest socioeconomic classes (28)

Seasonal variations in BP have immediate and long-term implications. Mortality from acute myocardial infarction and stroke and sudden cardiac arrest tend to be greater in winter months than in summer months. (41, 42). BP is a key factor for calculating the long-term risk of cardiovascular disease (43). An increase or decrease in BP due to seasonal variations may influence the calculation of CVD mortality risk using the well known Framingham risk score. For example, a subject screened in the winter, rather than in the summer, may have a higher estimated risk of CVD disease and, thus a higher likelihood of receiving a clinical preventive treatment such as a statin. Existing hypertension guidelines recommend BP monitoring every six months once a patient with hypertension is diagnosed and the underlying condition is well controlled with appropriate medication (44). It may be necessary to consider seasonal variations in BP measurements for CVD mortality risk scores and clinical guidelines.

Future research may focus on assessing the relationship between incidence of cardiovascular events (e.g. myocardial infraction or stroke) and seasonal blood pressure. This work may further assist clinicians with management of patients with higher cardiovascular risk(17). One study indicate how 2-5 mm Hg reduction in BP may lead to decrease in 3-7% in total mortality rate and 4-9% and 6-14% in mortality from coronary heart disease and stroke respectively.(45). Future research may also focus on BP measured in clinical setting measurements as compared to ambulatory BP monitoring or the self-BP measurements performed by individuals at home(14).

## Strength and limitations

The patient population in this study originated from routinely collected data from primary care clinics across the greater Toronto region. The overall patient demographics of this primary care repository are similar with respect to the Canadian census data(46). The clinical BP data in UTOPIAN primary care repository is further part of a single nationwide electronic medical record based chronic disease surveillance system.

There are limitations to the use of electronic medical records data in research. Some physicians may record blood pressure, weight and other such measurements in the text-based progress notes rather than in the designated fields; the data is not recognized as a BP if entered in these notes. The method of BP collection in routinely collected EMR data is unlikely to be standardized across practitioners. Many factors, including whether the patient was rested before the measurement, whether the correct cuff size was used, whether the physician or the nurse measured the blood pressure and whether a manual or automatic measurement device was used may influence BP measurements (32).

## Conclusion

In our real-world dataset, we found strong seasonal oscillations in blood pressure, especially among the elderly and patients with hypertension in this primary care population. Seasonal variations in blood pressure should be considered by primary care clinicians when optimizing strategies for regulating blood pressure and lowering cardiovascular risk for their patients.

## Data Availability

This work was supported by UTOPIAN, which is part of the Research and Advocacy Program at the Department of Family and Community Medicine, University of Toronto. The EMR database is not publicly available and requires appropriate credentials to access.

## Competing interest

None detected

## Funding

This work was supported by UTOPIAN, which is part of the Research and Advocacy Program at the Department of Family and Community Medicine, University of Toronto. Dr Greiver is supported through the Gordon F. Cheesbrough Research Chair in Family and Community Medicine from North York General Hospital. The opinions, results and conclusions reported in this paper are those of the authors and are independent from the funding sources.

## Acknowledgements

We are grateful to the physicians and patients who allow data sharing for UTOPIAN and for other Practice Based Research Networks contributing to the National Diabetes Repository. The results or views expressed are those of the author(s) and not necessarily those of UTOPIAN.

## Appendices

**Figure S1:**
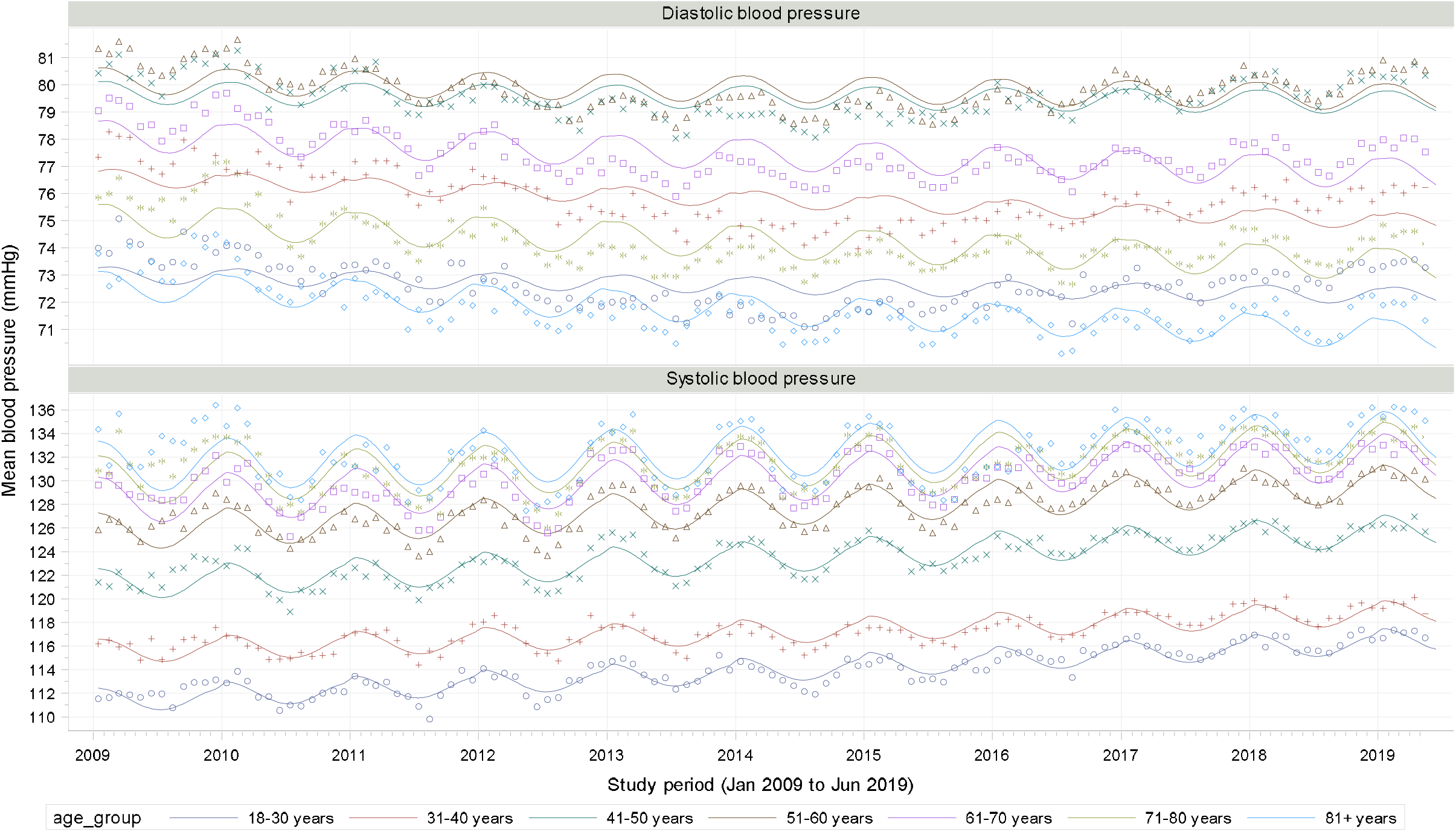
Sinusoidal curves for systolic and diastolic blood pressure by patient age groups.

**Figure S2:**
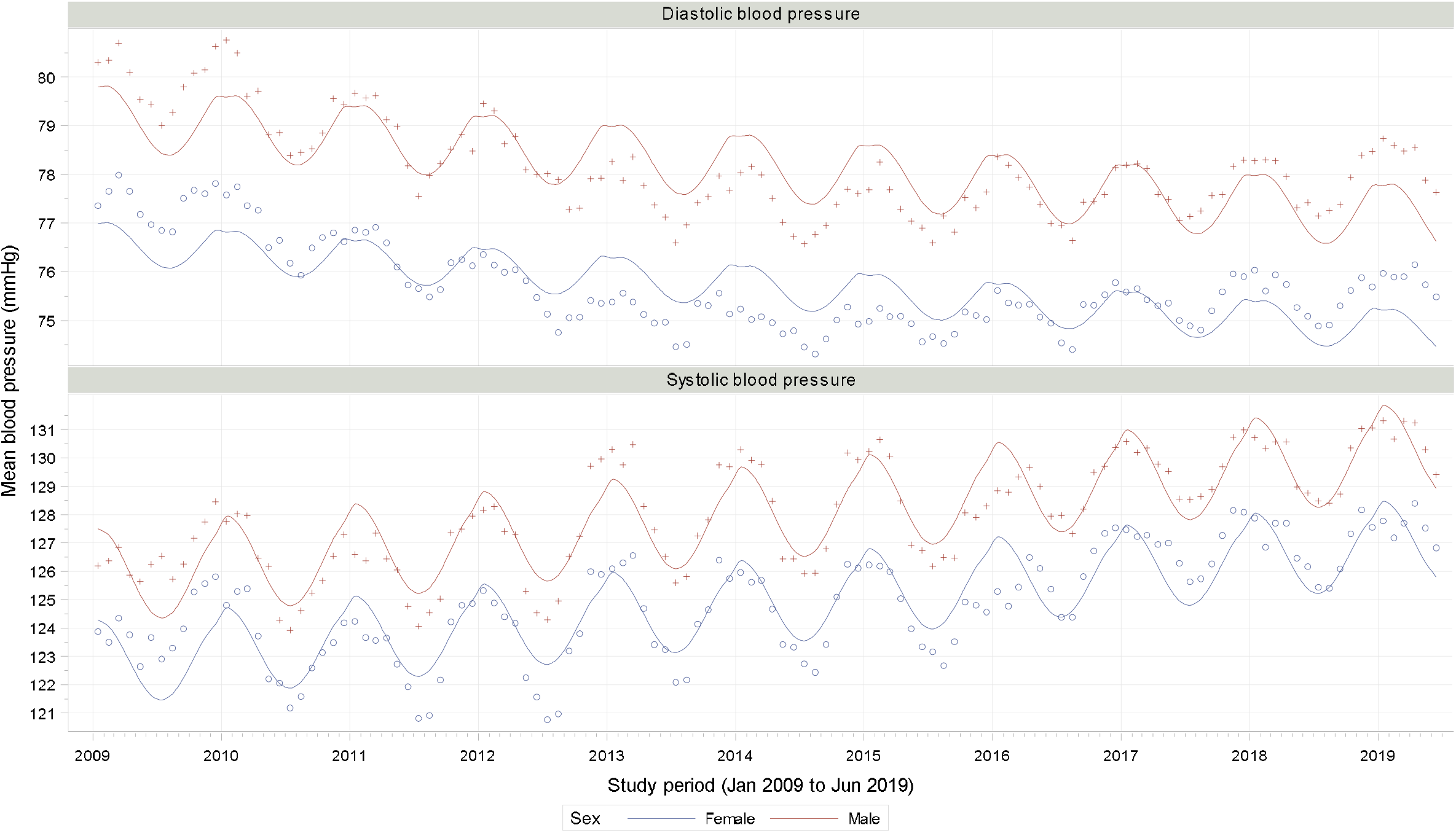
Sinusoidal curves for systolic and diastolic blood pressure by patient sex.

**Figure S3:**
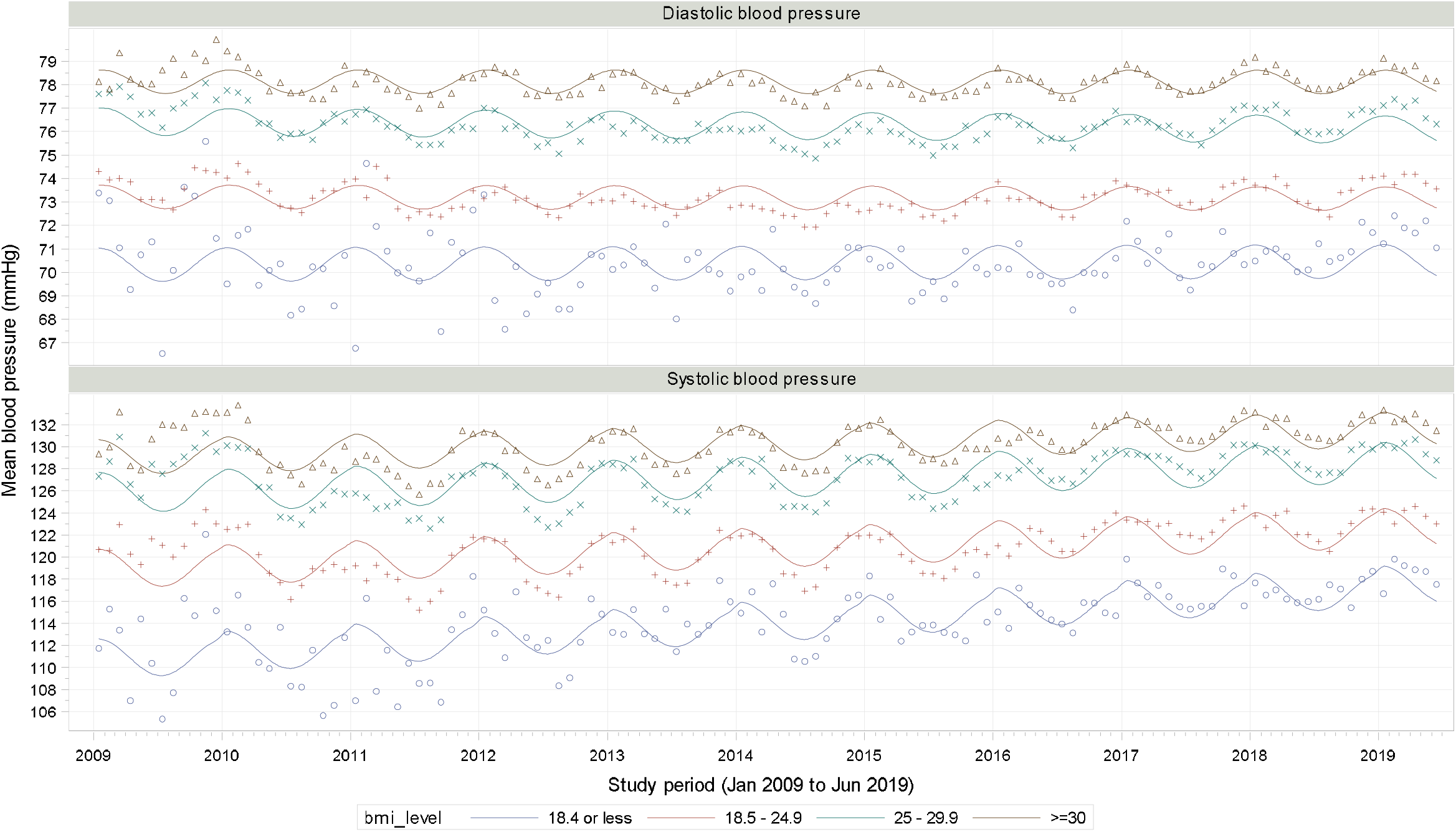
Sinusoidal curves for systolic and diastolic blood pressure by BMI groups.

**Figure S4:**
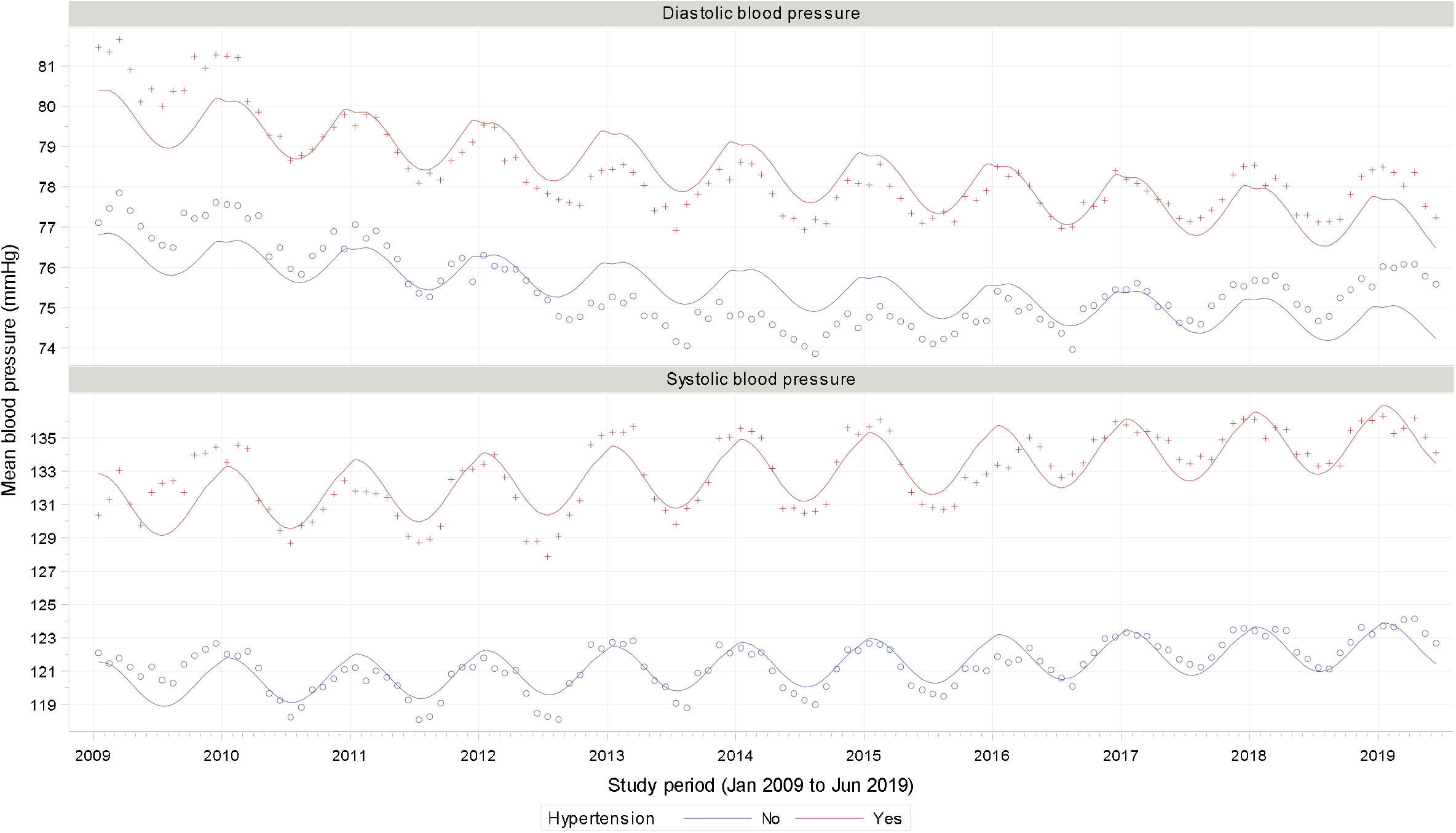
Sinusoidal curves for systolic and diastolic blood pressure by hypertensive status.

**Figure S5:**
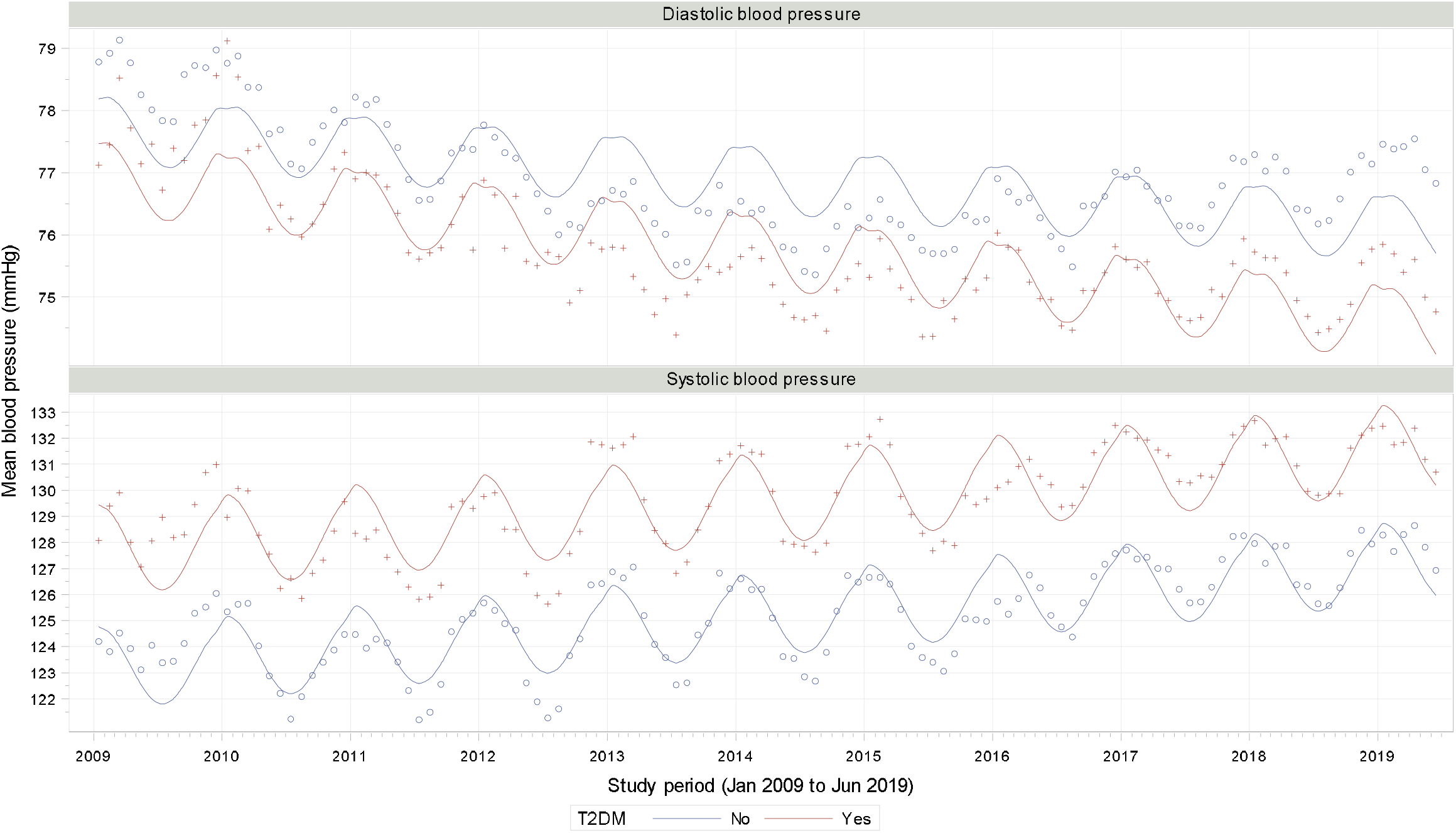
Sinusoidal curves for systolic and diastolic blood pressures by diabetic status.

**Figure S6:**
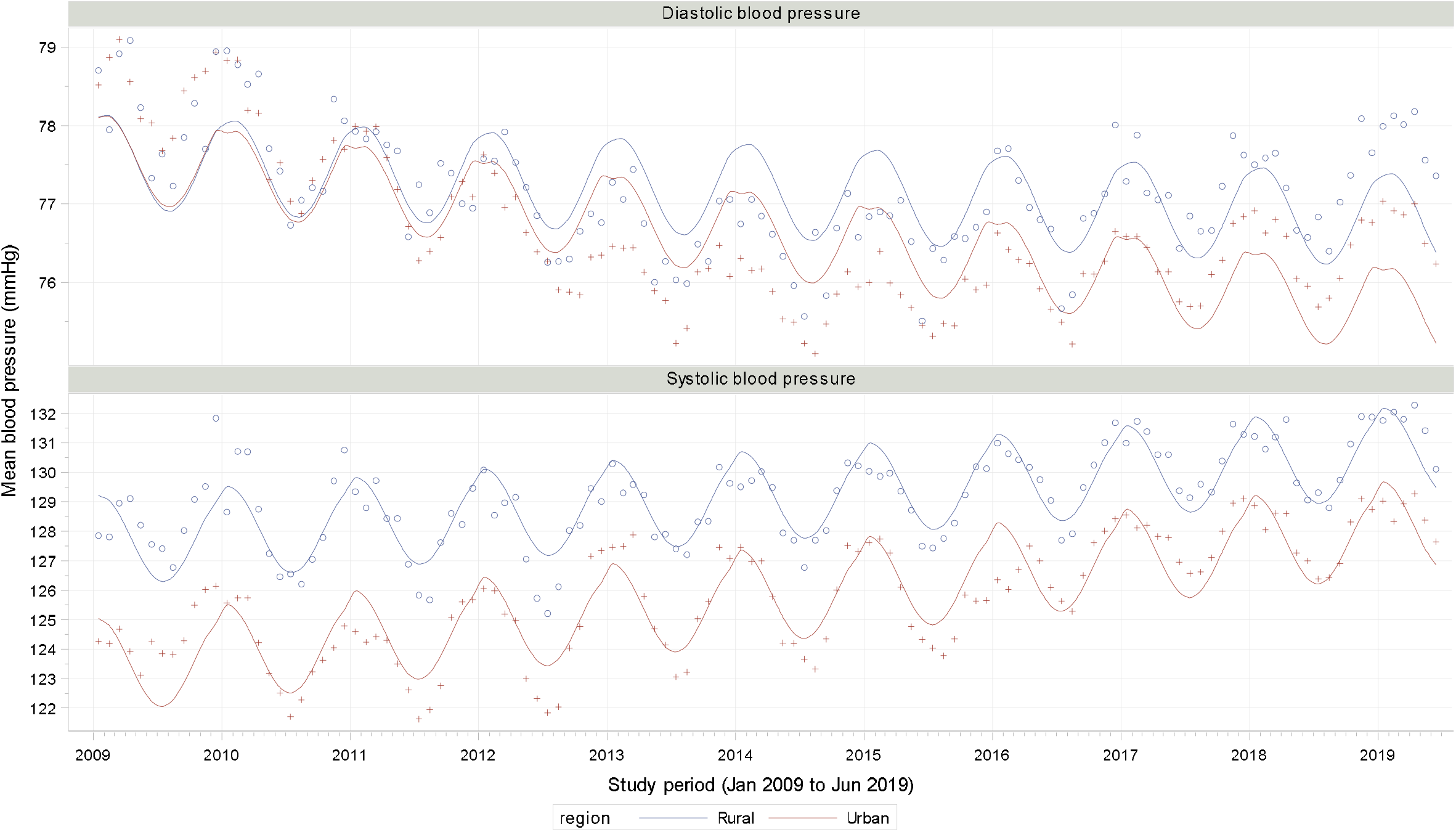
Sinusoidal curves for systolic and diastolic blood pressure by urban/rural region.

**Figure S7:**
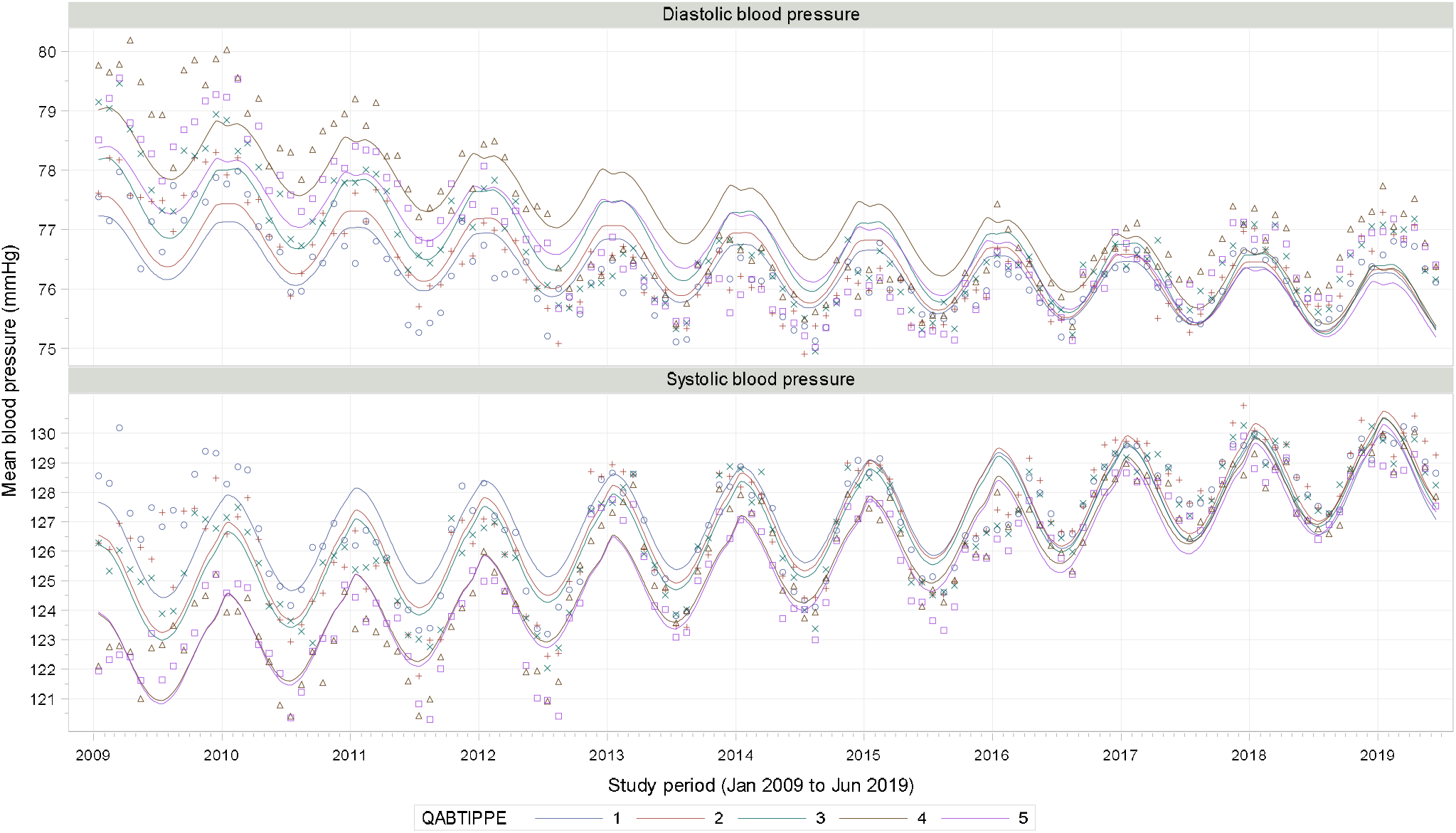
Sinusoidal curves for systolic and diastolic blood pressure by income quintiles.

## Notes

### Competing Interest Statement

The authors have declared no competing interest.

### Author Declarations

The study was reviewed and approved by the University of Toronto's Research Ethics Board.

